# The roles of stewardship and socioeconomic status in geographic variation in antibiotic prescribing: an observational study

**DOI:** 10.1101/2020.05.11.20098491

**Authors:** Stephen M Kissler, R Monina Klevens, Michael L Barnett, Yonatan H Grad

## Abstract

**Importance:** Reducing geographic disparities in antibiotic prescribing is a central public health priority to combat antibiotic resistance, but the drivers of this variation have been unclear.

**Objective:** To measure how differences in outpatient visit rates per 1,000 individuals (‘observed disease’) and in antibiotic prescribing rates per outpatient visit (‘stewardship’) each contributed to a gap in monthly antibiotic prescriptions per 1,000 individuals in Boston *vs*. the rest of Massachusetts, and to identify spatial correlates of the medical conditions that contributed most to the gap.

**Design:** We conducted an observational study of medical insurance claims collected between January 2011 and September 2015.

**Setting:** Outpatient visits in Massachusetts.

**Participants:** 5,028,609 Massachusetts residents under the age of 65 with at least one full month of primary medical insurance during the study period.

**Exposures:** Residence in Boston *vs*. the rest of Massachusetts.

**Main outcomes:** (1) The proportion of the gap in antibiotic prescribing rate between Boston and the rest of Massachusetts attributable to observed disease *vs*. stewardship for 20 medical conditions previously associated with antibiotic prescribing; and (2) geographic correlations between childhood vaccination, air pollution, density of children, and social deprivation index with the medical conditions that best explain the gap in prescribing.

**Results:** The average monthly antibiotic prescribing rate per 1,000 individuals in Boston was 51.8 (IQR 43.4, 57.5) *vs*. 65.2 (IQR 54.8, 74.5) in the rest of Massachusetts. Geographic differences in outpatient visit rates accounted for 56.0% of the gap in antibiotic prescribing between Boston and the rest of the state, while differences in stewardship accounted for just 19.8% of the gap. Geographic differences in outpatient visit rates for sinusitis, pharyngitis, and suppurative otitis media accounted for 43.3% of the gap in prescribing between Boston and the rest of Massachusetts. Social deprivation index best explains the geographic variation in outpatient visit rates for respiratory illness, with fewer visits in census tracts with higher social deprivation index.

**Conclusions and relevance:** Interventions aimed at reducing geographic disparities in antibiotic prescribing should target the drivers of outpatient visits for respiratory illness and should account for possible under-utilization of health services in areas with the lowest antibiotic consumption. Our findings challenge the conventional wisdom that prescribing practices are the main driver of geographic disparities in antibiotic use.

## Introduction

Antibiotic resistance poses a mounting threat to human health, with the number of deaths from resistant infections projected to reach up to 10 million by 2050^1^. Antibiotic resistance is driven by the consumption of antibiotics^2^, which varies geographically between countries^3^, states/provinces^4-6^, and local communities^7^. Reducing antibiotic prescribing is a central public health strategy to limit the emergence and spread of antibiotic resistance^8^. Two key variables govern antibiotic prescribing rates: the rate of seeking care in response to illness (*i.e*. the rate of ‘observed disease’), and the probability of receiving an antibiotic prescription once care is sought (*i.e*. ‘stewardship’). These in turn may be influenced by many other factors, including the incidence of illness, patient expectations^9^, accessibility of care^10^, physician age^11^, and provider type^6^. Measuring the relative roles of observed disease and stewardship in driving geographic disparities in antibiotic prescribing can point to the more specific mechanisms that may underlie those disparities^12^.

Much stewardship is focused on reducing inappropriate prescribing. Assessing the amount of inappropriate prescribing, though, is difficult, as it presumes estimates of the true incidence of infections that warrant treatment with antibiotics. Instead, a widely-accepted strategy for determining the target level of appropriate prescribing is to calibrate to the geographic locations with the lowest prescribing rates^13,14^. Still, low antibiotic prescribing rates do not necessarily imply successful antibiotic stewardship. They could instead reflect lower rates of disease or under-utilization of healthcare services due to social or economic barriers. To set appropriate goals for antibiotic prescribing and to choose the interventions most likely to attain them, it is critical to understand the drivers of geographic variation in antibiotic prescribing.

Large geographic variations in outpatient antibiotic prescribing have been noted at state^15,16^ and national levels^17,18^. In the United States, geographic variation in outpatient antibiotic prescribing is associated with the density and specialty of providers^6^ – pointing to a possible contribution from stewardship – but also with obesity, age, and outpatient visit rates for respiratory illness^6,19^, suggesting that the amount of observed disease may also vary. A direct comparison of stewardship and observed disease and their impact on the geography of antibiotic prescribing has not yet been possible due to the lack of representative, geographically resolved, individual-level data that allow antibiotic prescriptions to be linked with the outpatient visits that prompt them.

Massachusetts provides an ideal test case given that over 97% of residents have health insurance^20^, and the geographic variation in *per capita* antibiotic prescribing rate at the census tract level is of the same order as the geographic variation in antibiotic prescribing between US states^7^. Here, we analyzed a uniquely detailed medical insurance claims dataset from Massachusetts and quantified the degree to which age-stratified geographic differences in outpatient visit rates and antibiotic stewardship for 20 medical conditions explained a large gap in antibiotic prescribing between Boston and the rest of Massachusetts^7^. For the medical conditions that contributed most to the geographic disparities in antibiotic prescribing, we assessed correlations with childhood vaccination, air pollution, density of children, and social deprivation index (SDI).

## Methods

### Study sample and data sources

Medical and pharmacy claims data were extracted from the Massachusetts All-Payer Claims Database (APCD), 5^th^ edition^21^. The APCD contains insurance claims filed between January 1^st^, 2011 and September 30^th^, 2015 for 94% of Massachusetts residents (*n =* 5,028,609) under the age of 65 (**Table** 1). We excluded claims associated with individuals over the age of 65, since most of these individuals use traditional fee-for-service Medicare for their primary health insurance, and fee-for-service Medicare claims are not included in the APCD. We also excluded claims filed after October 1^st^, 2015 when the ICD-10 diagnosis classification system was adopted nationally, unpredictably shifting the observed prevalence of many conditions due to differences in coding^22^. We extracted all pharmacy claims for antibiotics (*n* = 17,680,795) using previously described methods^7^. Each antibiotic claim was associated with a patient identifier, the patient’s age, the patient’s census tract, and the fill date^7^.

Outpatient physician encounters (‘visits’) were extracted from the APCD on the basis of Current Procedural Terminology (CPT) codes associated with outpatient procedures (*n* = 81,807,743). Each visit was associated with a patient identifier, the patient’s age, the patient’s census tract, the service date, and up to seven ICD-9 coded diagnoses.

Geographical boundaries for census tracts were obtained from the US Census Bureau’s TIGER shapefiles^23^. We defined ‘Boston’ as the census tracts associated with the US Census Bureau’s designated place of Boston City^24^. The fraction of the population under the age of 10 was obtained from the 2015 US Census Bureau’s American Fact Finder^24,25^. The proportion of students meeting school requirements for immunizations was obtained at the county level from the Massachusetts Department of Public Health^26^. Particulate matter concentrations under 2.5 micrometers (PM2.5) were obtained from the Centers for Disease Control and Prevention^27^. We used the social deprivation index (SDI),^28^ an index that aggregates statistics related to poverty, education, single-parenthood, housing type, and employment, as a measure of socioeconomic variables relevant to healthcare access.

### Defining medical conditions and associated prescriptions

Each diagnosis in our study sample was mapped to one of 20 medical conditions previously defined as associated with antibiotic prescribing^13^. For each visit, the diagnosis that ranked highest on the scale of antibiotic appropriateness^13^ (Tier 1: always appropriate; Tier 2: sometimes appropriate; Tier 3: never appropriate) was assigned as that visit’s primary diagnosis. We linked antibiotic prescriptions with visits on the basis of patient identifier and time between visits and prescription events^29,30^. Each prescription could be linked to at most one visit; if a patient underwent multiple visits within the week preceding a filled antibiotic prescription, only the visit nearest to the prescription was linked. A prescription could fail to link with a visit if the associated visit did not qualify as an outpatient physician visit (e.g. the prescription was written during a dentistry visit or as a follow-up from a hospital stay), if the patient received the antibiotic without a formal outpatient visit (i.e., ‘phantom prescribing’^31^), if the prescription was filled more than one week after the visit, or if the patient used separate forms of insurance for the visit and the prescription fill, thereby associating the two claims with different patient IDs.

### Study outcomes and covariates

Our primary outcomes were (a) the difference in antibiotic prescribing rate per 1,000 individuals between Boston and the rest of Massachusetts, as well as the proportion of this gap attributable to geographic differences in (b) outpatient visit rates per 1,000 individuals and (c) the probability of receiving an antibiotic prescription given an outpatient visit. We considered age, gender, vaccination rates, air pollution, and SDI as covariates.

### Calculating relative risks

We used Poisson regression to estimate the age- and sex-adjusted relative risk of (a) undergoing an outpatient visit and (b) receiving an antibiotic given an outpatient visit in a given month for individuals within Boston *vs*. outside of Boston, for the 20 medical conditions.

### Decomposing the geographic gap in prescribing

We calculated the proportion of the gap in antibiotic prescriptions per 1,000 individuals dispensed in Boston *vs*. the rest of the state that was attributable to (a) geographic differences in outpatient visit rates per 1,000 individuals and (b) geographic differences in prescriptions given per outpatient visit (*i.e*. stewardship) for the 20 medical conditions, stratified by age decile. To do so, we first expressed the total number of antibiotic prescriptions per 1,000 individuals in Boston (*R*) and in the rest of Massachusetts 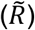 as the sum of the number of prescriptions *r_j_* attributable to each medical condition *j* plus the number of prescriptions *u* that were not linked to an outpatient visit:

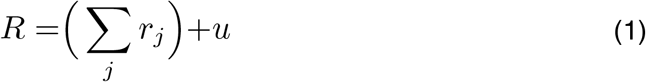

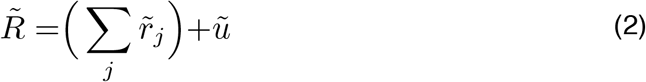

Throughout, the tilde (~) denotes ‘the rest of Massachusetts’, *i.e*. not Boston. Next, we expressed the condition-specific prescribing rates per 1,000 individuals (*r_j_* and *u*) as the sum of the prescribing rate per 1,000 individuals attributable to each age decile *a*:

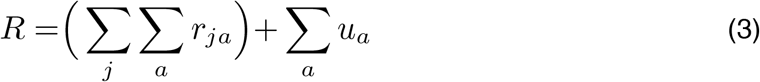

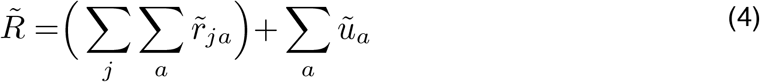

We then expressed each condition- and age-specific prescribing rate *r_ja_* as the product of the outpatient visit rate per 1,000 individuals for that condition and age decile (*v_ja_*) and the probability of receiving an antibiotic prescription at a visit for that condition (*p_ja_*):

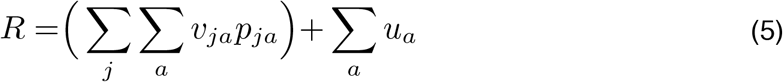

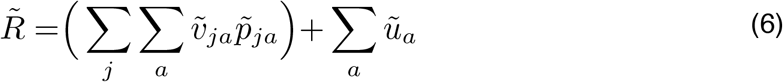

Subtracting *R* from 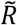 gives the gap in antibiotic prescribing rate per 1,000 individuals between Boston and the rest of Massachusetts:

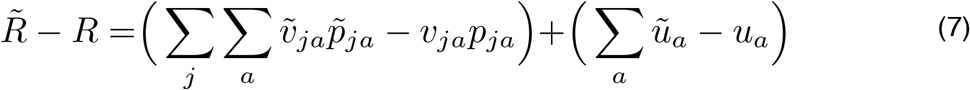

Finally, we separated the Boston *vs*. non-Boston difference in prescribing rate per 1,000 individuals for each condition and age decile into components attributable to different outpatient visit rates per 1,000 individuals (observed disease), different prescribing rates per outpatient visit (stewardship), and a residual:

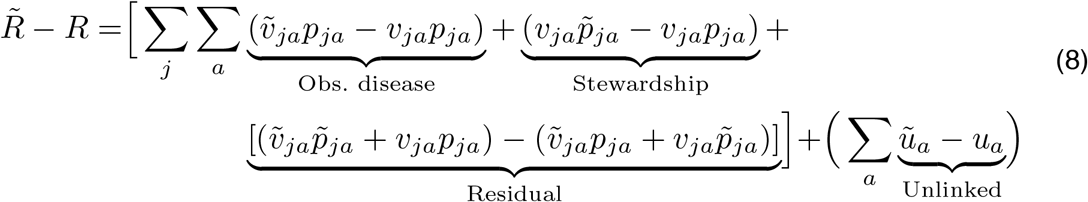

The ‘observed disease’ term measures the number of antibiotic prescriptions per 1,000 individuals that would have been given in the rest of Massachusetts if only the outpatient visit rate (*v_j_*_a_), but not the probability of receiving a prescription (*p_ja_*), had differed from the outpatient visit rate in Boston. The ‘stewardship’ term is interpreted similarly. The ‘residual’ term is the quantity needed to make Eq. 8 equal to Eq. 7; this represents a synergistic (non-additive) effect between outpatient visits and stewardship not directly attributable to either mechanism. To calculate the proportion of the gap in prescribing attributable to observed disease *vs*. stewardship (as well as the residual and unlinked prescriptions) by medical condition and age decile, we divided each bracketed term by the total size of the gap, 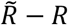.

### Identifying correlates of outpatient visit rates

We used Poisson regression to test whether air pollution, fraction of the population under the age of 10, fraction of kindergarteners meeting school vaccine requirements, and/or SDI predicted geographic variation in outpatient visit rates for the medical conditions that contributed most to the gap in antibiotic prescribing, adjusting for age and sex. We calculated the regressions across all census tracts in Massachusetts (including Boston) and also separately for Boston census tracts to examine whether correlates of prescribing at the state level were retained at the city level.

#### Results

Between January 2011 and September 2015, the average monthly antibiotic prescribing rate per 1,000 individuals in Boston was 51.8 (IQR 43.4, 57.5) *vs*. 65.2 (IQR 54.8, 74.5) in the rest of Massachusetts. **Figure 1A** depicts the geographic variation in antibiotic prescribing rates per 1,000 individuals in Massachusetts by census tract, illustrating the relatively low antibiotic prescribing rates in Boston and in western Massachusetts. Similar geographic patterns governed the outpatient visit rates per 1,000 individuals for Tier 1 and 2 conditions (**Figure 1B**) and the probability of receiving an antibiotic prescription given a Tier 1 or 2 outpatient visit **(Figure 1C**). Despite a decline in the overall antibiotic prescribing rate in Massachusetts over the study period^7^, the geographic variation in antibiotic prescribing remained similar from year to year (**Figures S1-5**).

**Figure 1.**
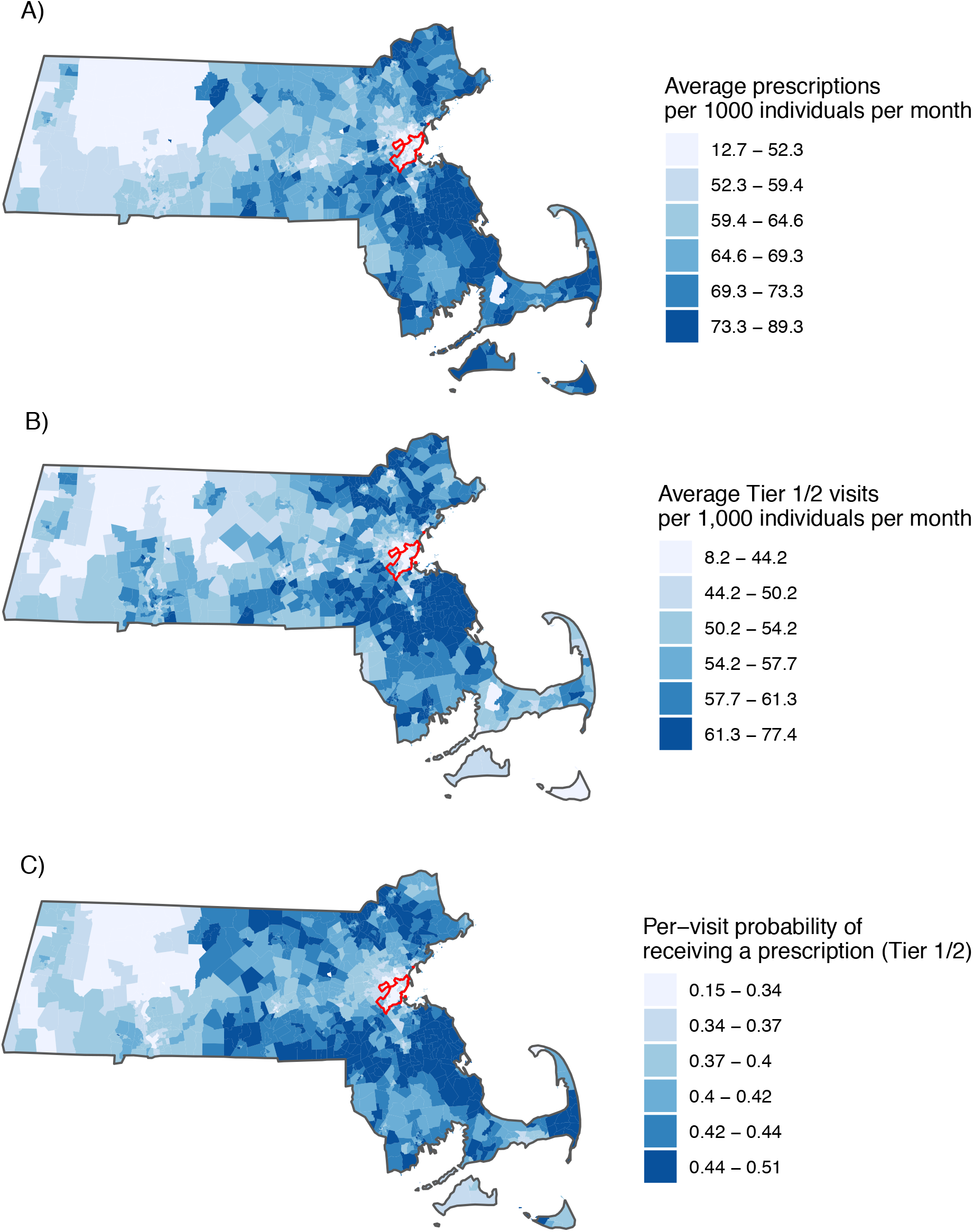
(A) Mean number of monthly antibiotic prescriptions per 1,000 individuals in Massachusetts by census tract. (B) Mean number of monthly Tier 1 and 2 outpatient visits per 1,000 individuals in Massachusetts by census tract. (C) Mean monthly probability of receiving an antibiotic prescription given an outpatient visit for a Tier 1 or 2 condition by census tract. Boston is outlined in red. Cutoffs for the color scales are sextiles.

The relative risk of undergoing an outpatient visit for a respiratory condition, adjusted for age and sex, ranged from 1.17 (95% CI 1.16, 1.18) to 3.12 (95% CI 2.69, 3.61) times higher in census tracts outside of Boston than within Boston, depending upon the condition (**Figure 2A**). For non-respiratory conditions the relative risk of an outpatient visit was similar between Boston and the rest of the state (**Figure 2A**). The relative risk of receiving an antibiotic prescription given an outpatient visit outside of Boston *vs*. within Boston ranged from 0.91 (95% CI 0.90, 0.92) for “other genitourinary conditions” to 1.49 (95% CI 1.46, 1.51) for viral upper respiratory infections (**Figure 2B**).

**Figure 2.**
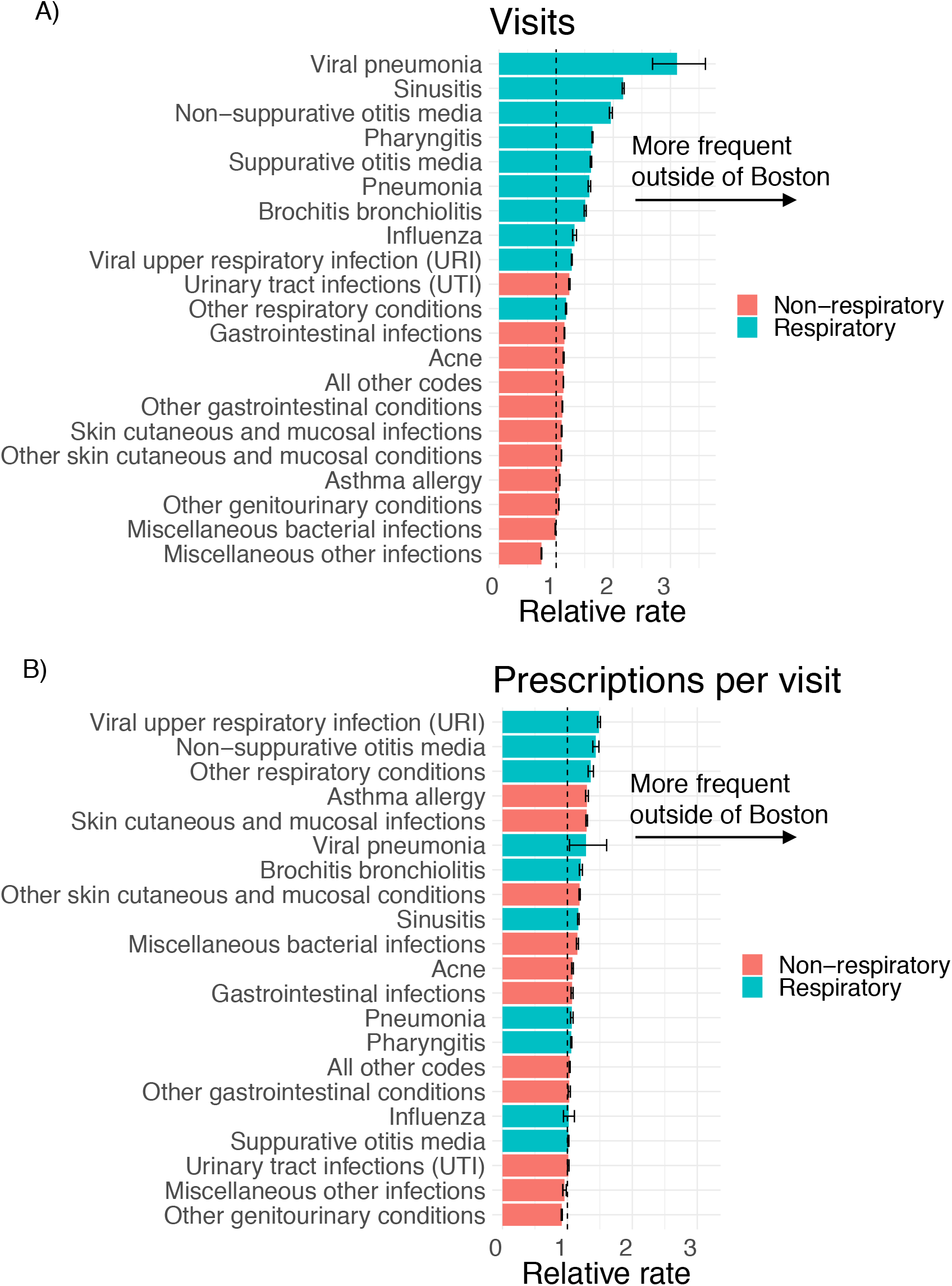
(A) Age- and sex-adjusted relative rate of undergoing an outpatient visit in a given month in the rest of Massachusetts relative to Boston by medical condition. (B) Age- and sex-adjusted relative rate of receiving an antibiotic prescription at a given outpatient visit in the rest of Massachusetts relative to Boston by medical condition. The dashed line marks no difference in risk.

**Table 1.** Characteristics of the study population. Boston City is contained fully within Suffolk County.

| **Demographic** | **Number** | **Percent** |
| --- | --- | --- |
| **Total** | 6,195,517 | 100 |
| **Age** |  |  |
| 0-2 | 513,457 | 8.29 |
| 3-5 | 245,230 | 3.96 |
| 6-9 | 314,867 | 5.08 |
| 10-14 | 397,840 | 6.42 |
| 15-19 | 466,534 | 7.53 |
| 20-29 | 1,173,525 | 18.9 |
| 30-39 | 933,589 | 15.1 |
| 40-49 | 926,482 | 15.0 |
| 50-59 | 879,366 | 14.2 |
| 60-64 | 344,627 | 5.56 |
| **Female** | 3,142,441 | 50.7 |
| **County** |  |  |
| Barnstable | 170,119 | 2.75 |
| Berkshire | 110,172 | 1.78 |
| Bristol | 477,628 | 7.71 |
| Dukes | 14,286 | 0.231 |
| Essex | 710,469 | 11.5 |
| Franklin | 60,694 | 0.980 |
| Hampden | 431,392 | 6.96 |
| Hampshire | 117,954 | 1.90 |
| Middlesex | 1,493,939 | 24.1 |
| Nantucket | 11,848 | 0.191 |
| Norfolk | 630,829 | 10.2 |
| Plymouth | 441,639 | 7.13 |
| Suffolk | 778,107 | 12.6 |
| Worcester | 746,441 | 12.0 |
| **Boston City** | 664,960 | 10.7 |

**Table 2.**
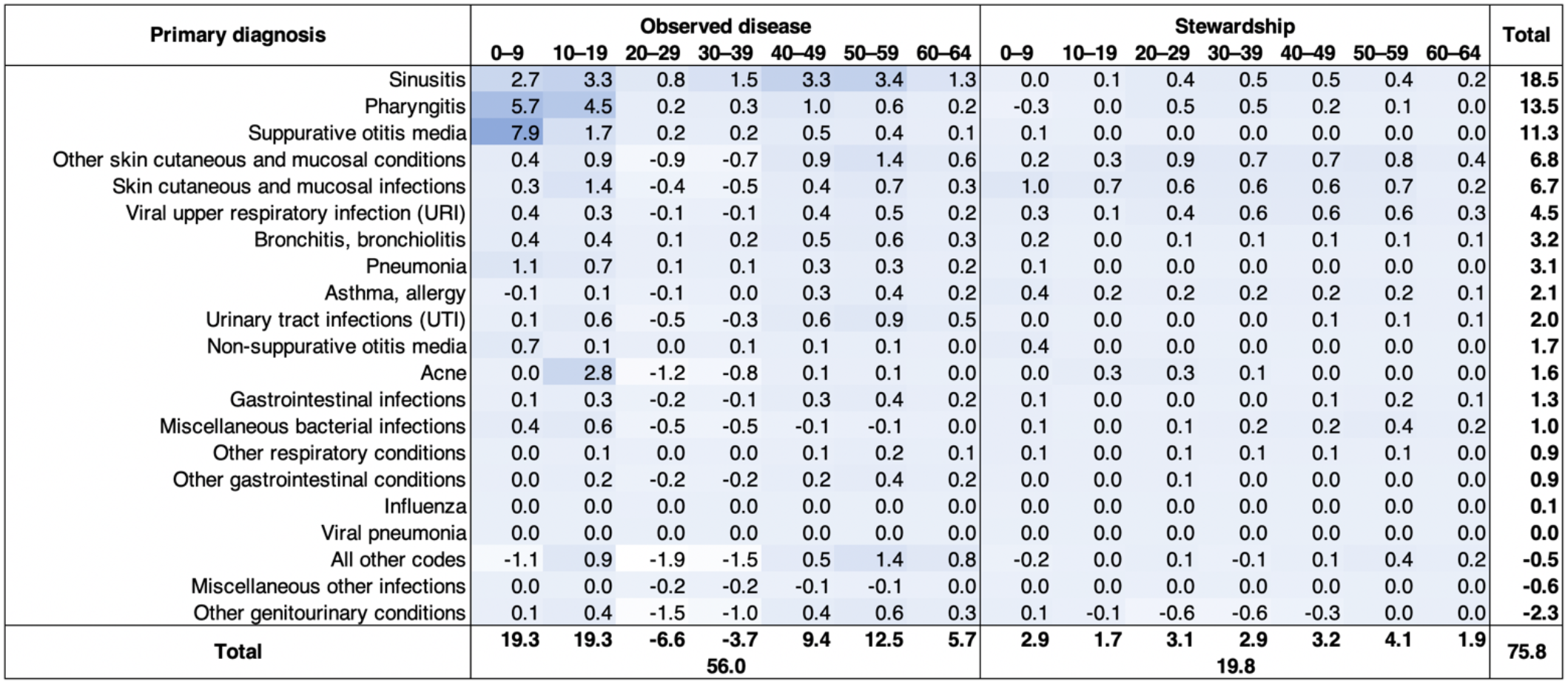
Percent contribution of geographic differences in outpatient visit rates (observed disease) and per-visit prescribing rate (stewardship) to the gap in antibiotic prescribing rate per 1,000 individuals between Boston and the rest of Massachusetts, by medical condition and age group. Higher percentages are denoted by more intense blue. Marginal sums, also in units of percent contribution to the gap, are listed along the bottom and left. Negative values indicate that the outpatient visit rate per 1,000 individuals and/or the probability of receiving an antibiotic prescription given an outpatient visit was higher in Boston than in the rest of Massachusetts, in opposition of the prevailing trend. In total, 75.8% of the gap is attributable to differences in outpatient visit rates and per-visit prescribing rates; the rest (24.2%) is attributable to unlinked prescriptions and non-additive effects.

**Table 3.** Sex- and age-adjusted Poisson regression coefficients and 95% confidence intervals for four predictors of outpatient visit rates per 1,000 individuals at the census tract level and the three medical conditions that contributed most to the gap in prescribing between Boston and the rest of Massachusetts.

|  | **Sinusitis** | **Pharyngitis** | **Suppurative otitis media** |
| --- | --- | --- | --- |
| **Fraction of kindergarteners fully vaccinated** | 0.0340<br>(0.0336, 0.0343) | 0.0268<br>(0.0265, 0.0271) | 0.0236<br>(0.0232, 0.0239) |
| **Fraction of population under 10** | -1.40<br>(-1.45, -1.35) | -0.545<br>(-0.582, -0.508) | -1.36<br>(-1.41, -1.31) |
| **Average PM2.5** | -0.310<br>(-0.313, -0.307) | -0.132<br>(-0.134, -0.129) | -0.272<br>(-0.275, -0.269) |
| **Social deprivation index** | -0.709<br>(-0.714, -0.703) | -0.621<br>(-0.625, -0.617) | -0.647<br>(-0.652, -0.641) |

In an age-stratified analysis, geographic differences in outpatient visit rates accounted for 56.0% of the gap in antibiotic prescribing between Boston and the rest of the state, while differences in stewardship accounted for just 19.8% of the gap (**Table 2**). The remaining 24.2% of the gap could not be definitively linked to either outpatient visit rates or stewardship differences; this quantity was attributable to residual non-additive effects between outpatient visit rates and stewardship (6.9% of the gap, see Eq. 8) and to prescriptions that could not be linked to an outpatient visit (17.3% of the gap). Geographic differences in outpatient visit rates for sinusitis, pharyngitis, and suppurative otitis media accounted for 43.3% of the geographic gap in antibiotic prescribing, a greater contribution than the other 17 conditions combined (32.5%). The contributions from pharyngitis and otitis media were mostly attributable to lower outpatient visit rates in Boston for children under the age of 10, while the contribution from sinusitis-related outpatient visits was spread more evenly across age groups (**Table 2**).

The rates of outpatient visits per 1,000 individuals for sinusitis, pharyngitis, and suppurative otitis media were all positively associated with the fraction of kindergarteners fully meeting school vaccine requirements, and were negatively associated with fraction of the population under the age of 10, average particulate matter (PM2.5) concentrations, and SDI, according to age- and sex-adjusted Poisson regression at the census tract level both across Massachusetts (**Table** 3) and within Boston (**Table S1**).

#### Discussion

The lower antibiotic prescribing rate in Boston *vs*. the rest of Massachusetts in 2011-2015 was primarily due to lower rates of outpatient visits in Boston *vs*. the rest of the state, especially for sinusitis, pharyngitis, and suppurative otitis media. The probability of receiving an antibiotic prescription given an outpatient visit was also lower in Boston than in the rest of Massachusetts, but the disparity in outpatient visit rates contributed substantially more than differences in stewardship to the geographic gap in antibiotic prescribing (56.0% *vs*. 19.8%). Children under the age of 10 contributed the most to the geographic gap in antibiotic prescribing. Social determinants of health, as measured by the SDI, provide the best explanation for the variation in outpatient respiratory illness visit rates observed across Massachusetts, with fewer outpatient visits in census tracts with higher SDI (*i.e*. lower socioeconomic status). All of the other correlations we assessed ran counter to intuition, probably due to confounding with factors that contribute to increased SDI. For example, air pollution is highest in areas with high SDI and vaccination is the lowest in these areas, making it appear as though high pollution and low vaccination predict low respiratory outpatient visit rates. The strong negative relationship between SDI and outpatient respiratory illness visit rates held within Boston as well, such that individuals from wealthier census tracts in Boston had relatively more outpatient visits – and received more antibiotics – than their immediate neighbors in poorer areas. Taken together, our findings suggest that the lower antibiotic prescribing rates in Boston *vs*. the rest of Massachusetts were not primarily due to better prescribing by Boston physicians, but rather to reduced healthcare access in the most impoverished areas of Massachusetts^32^ and/or an inflated tendency to seek medical care for respiratory illness in wealthier areas. An analysis of hospitalizations associated with respiratory illness by SDI might serve to evaluate this finding and assess whether fewer visits among the poor resulted in more complications.

Our study expands upon others that have identified correlates of geographic variation in antibiotic prescribing in the United States^6,7,19^, strengthening the evidence base linking antibiotic prescribing with outpatient respiratory visits^19^ and attributing that variation in part to socioeconomic status. Given the high level of access to care in Massachusetts, future studies should explore collaborations between public health and healthcare providers to address barriers to care that persist for some parts of the population. Our study complements others that have associated longitudinal declines in antibiotic prescribing with reductions in outpatient visits for respiratory illness^12,33^. Additionally, our findings underscore the utility of the SDI as a measure of reduced healthcare access and associated downstream effects^28^.

Our study was subject to a number of limitations. First, this is a cross-sectional observational study that does not establish causality. Confounding between antibiotic prescribing, stewardship and unobserved population characteristics could explain our findings beyond the factors we examined. Our sample did not include individuals over the age of 65, who are known to receive a high volume of antibiotic prescriptions^34^. Geographic patterns in antibiotic prescribing are consistent across age groups at the level of US states^6^, however, so we anticipate that the geographic patterns described here would remain similar if this age group were included. Also, our study period concluded in 2015, but since the geographic patterns in antibiotic prescribing remained consistent across years^7^, our findings likely remain applicable. Our sample was also limited to a single US state; different geographic drivers of prescribing may dominate at different geographic scales. However, because of the high insurance coverage and availability of medical services in Massachusetts, it is an ideal state to describe antibiotic prescribing. At the level of US states, the association between SDI and antibiotic prescribing appears reversed, with high SDI and high antibiotic prescribing rates coinciding in the southeast^15^ (see also **Figure S6**). Nevertheless, antibiotic prescribing remains strongly correlated with outpatient visits for respiratory illness at the national scale^19^, suggesting that the drivers of respiratory illness are also key drivers of antibiotic prescribing across geographic scales. We focused only on outpatient antibiotic prescriptions; however, these account for 80-90% of all antibiotic prescriptions^35^, so we believe we have captured the majority of prescriptions given in Massachusetts during the study period. We also did not capture antibiotics that may have been obtained without a prescription or those paid out of pocket, though we expect these are rare in a state with near-universal health insurance coverage.

Our findings challenge the conventional wisdom that prescribing practices are the main driver of geographic disparities in antibiotic use. This has implications for planning and evaluating interventions intended to reduce antibiotic consumption: interventions should address the drivers of outpatient visits for respiratory illness, and targets for antibiotic prescribing should be adjusted for potential under-utilization of healthcare services in the areas that use the fewest antibiotics. While outpatient visit rates for respiratory illnesses were strongly associated with SDI, other drivers, such as patient attitudes toward antibiotic prescribing and the incidence of actual illness, may exist. Further studies are needed to understand the relative importance of these factors, which will help to maximize the effectiveness of interventions.

#### Conclusions

Geographic disparities in antibiotic prescribing are driven largely by variable outpatient visit rates for respiratory illnesses, especially among children. In turn, outpatient visit rates for respiratory illness are lowest in settings with low socioeconomic status as measured by the SDI. Interventions aimed at reducing geographic disparities in antibiotic prescribing should target the drivers of outpatient visits for respiratory illness and should account for possible under-utilization of health services in areas with the lowest antibiotic consumption.

#### Author contributions

SMK assisted in conceptualization, performed formal analysis, and wrote the manuscript. RMK assisted in conceptualization, curated data, and edited the manuscript. MLB assisted in conceptualization, contributed methodology, and edited the manuscript. YHG assisted in conceptualization, contributed methodology, supervised the work, and edited the manuscript.

#### Exclusive license

The Corresponding Author has the right to grant on behalf of all authors and does grant on behalf of all authors, an exclusive license (or non-exclusive for government employees) on a worldwide basis to the BMJ Publishing Group Ltd to permit this article (if accepted) to be published in BMJ editions and any other BMJPGL products and sublicences such use and exploit all subsidiary rights, as set out in our license.

#### Competing interest statement

All authors have completed the Unified Competing Interest form (available on request from the corresponding author) and declare: no support from any organization for the submitted; no financial relationships with any organizations that might have an interest in the submitted work in the previous three years, no other relationships or activities that could appear to have influenced the submitted work.

#### Transparency declaration

The lead author (the manuscript’s guarantor) affirms that this manuscript is an honest, accurate, and transparent account of the study being reported; that no important aspects of the study have been omitted; and that any discrepancies from the study as planned have been explained.

#### Ethics statement

This study was a secondary analysis of data collected for other purposes and was not considered human subjects research and not reviewed by IRB.

#### Funding statement

No funding was received for conducting this work. The authors were employees of the Harvard Chan School of Public Health or Massachusetts Department of Public Health.

#### Patient and public involvement

No patients or members of the public were involved in the design, or conduct, or reporting, or dissemination plans of our research.

#### Data sharing statement

Massachusetts APCD data are available from the Center for Health Information and Analysis^21^.

## Data Availability

Data are available publicly from https://www.chiamass.gov/

## Supplemental information

**Figure S1.**
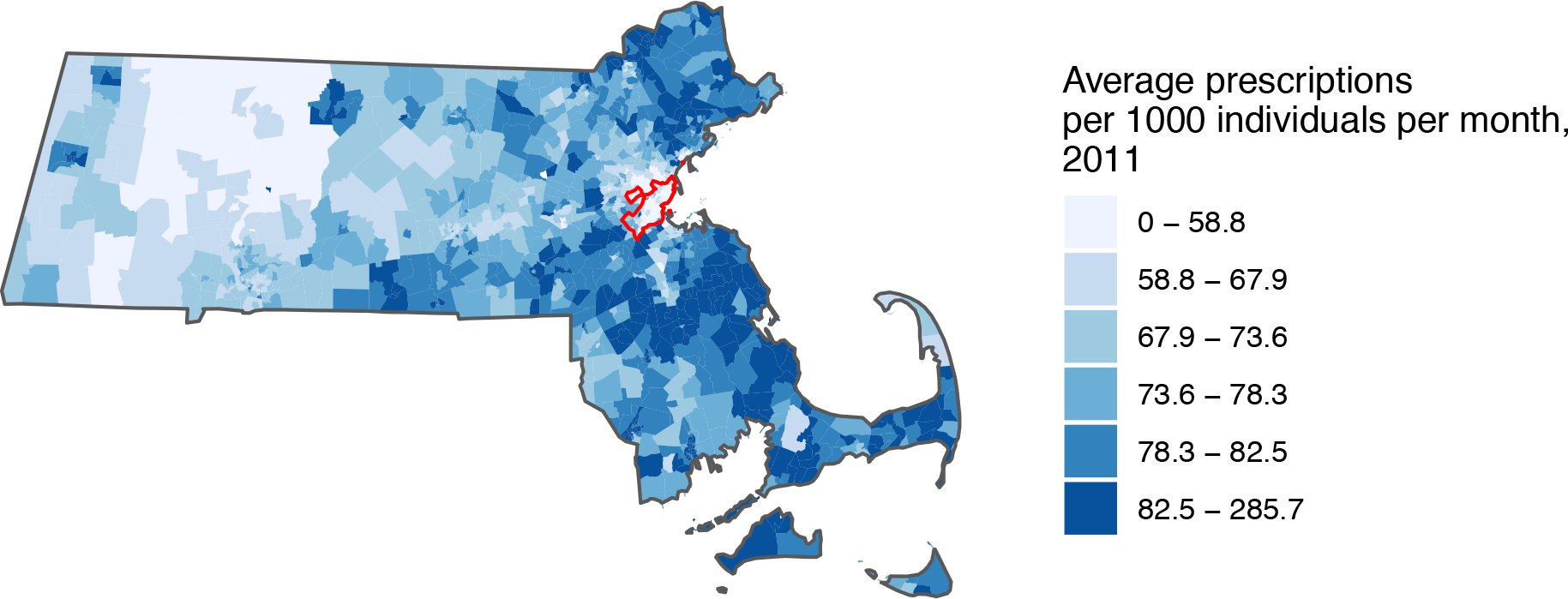
Average monthly antibiotic prescribing rate per 1,000 individuals by Massachusetts census tract in 2011.

**Figure S2.**
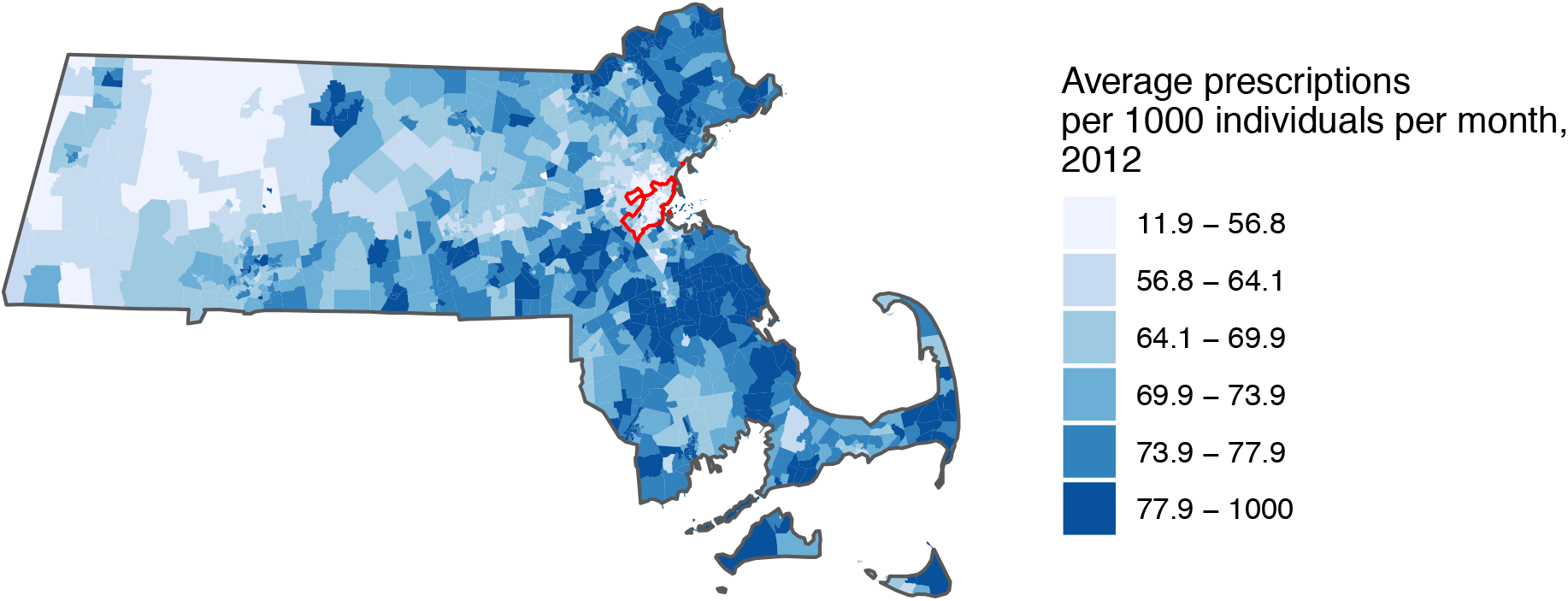
Average monthly antibiotic prescribing rate per 1,000 individuals by Massachusetts census tract in 2012.

**Figure S3.**
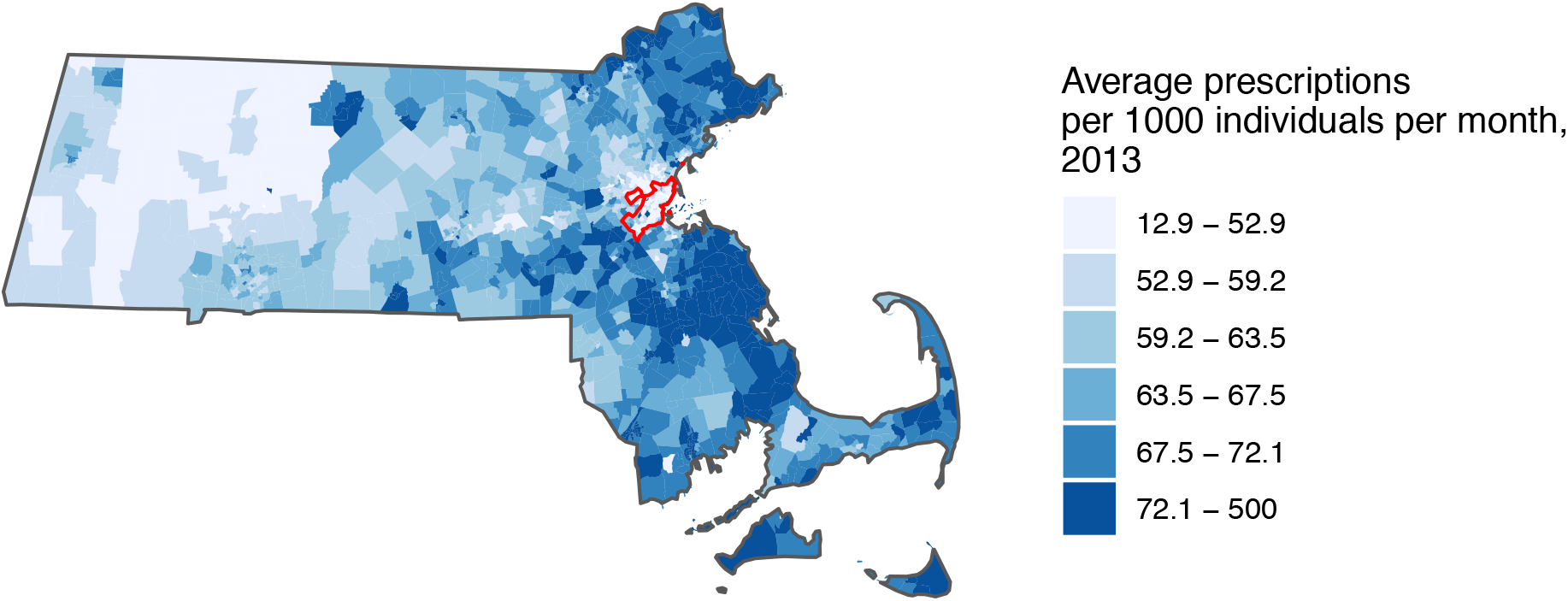
Average monthly antibiotic prescribing rate per 1,000 individuals by Massachusetts census tract in 2013.

**Figure S4.**
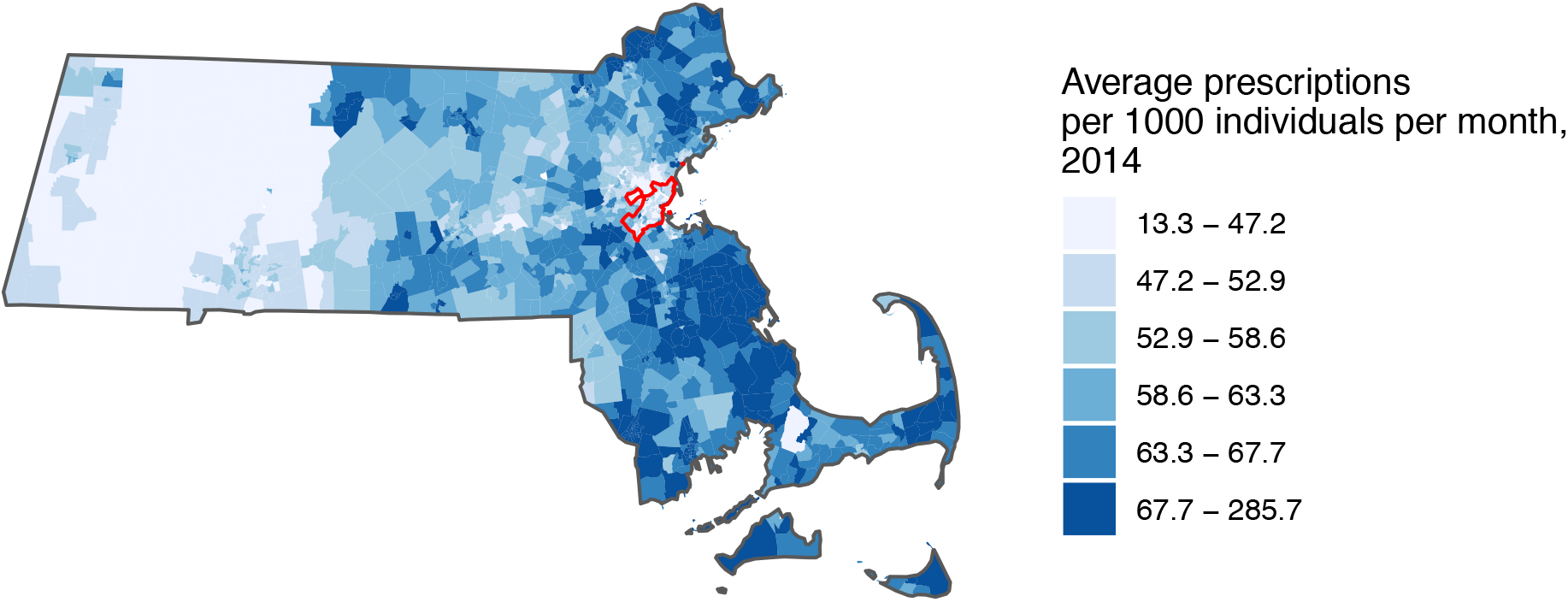
Average monthly antibiotic prescribing rate per 1,000 individuals by Massachusetts census tract in 2014.

**Figure S5.**
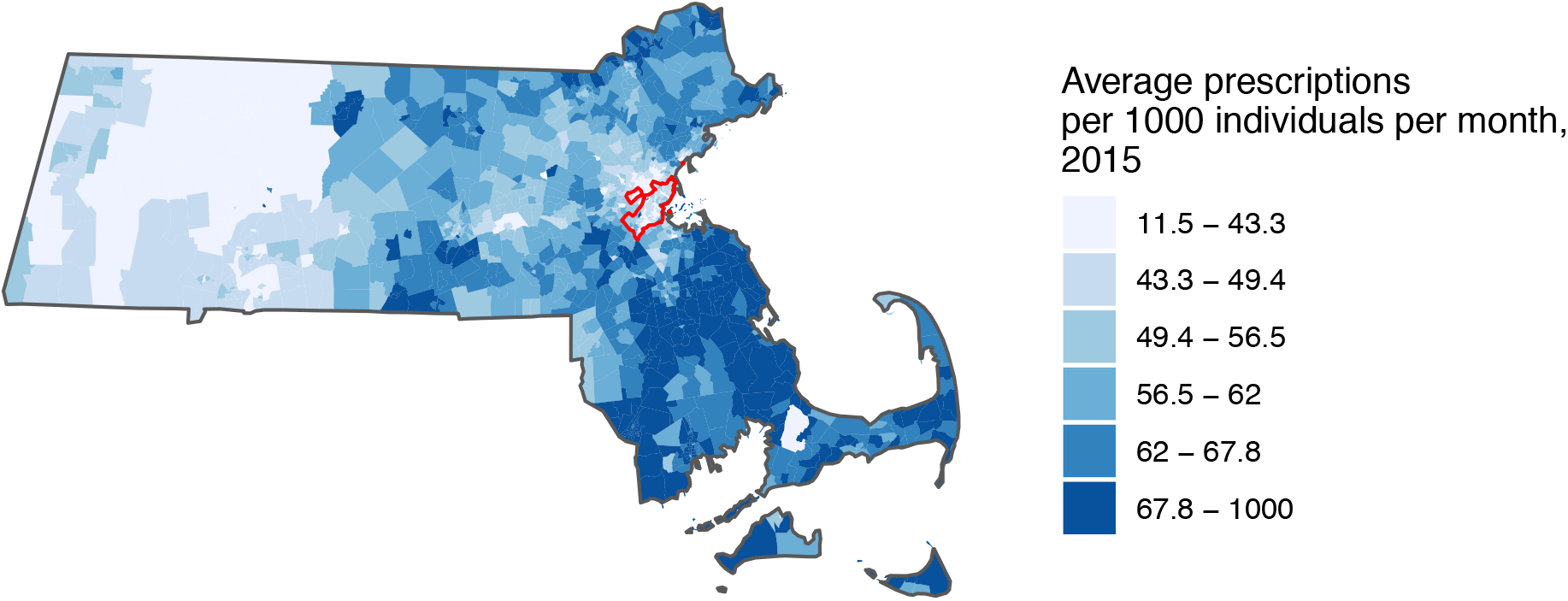
Average monthly antibiotic prescribing rate per 1,000 individuals by Massachusetts census tract in 2015.

**Figure S6.**
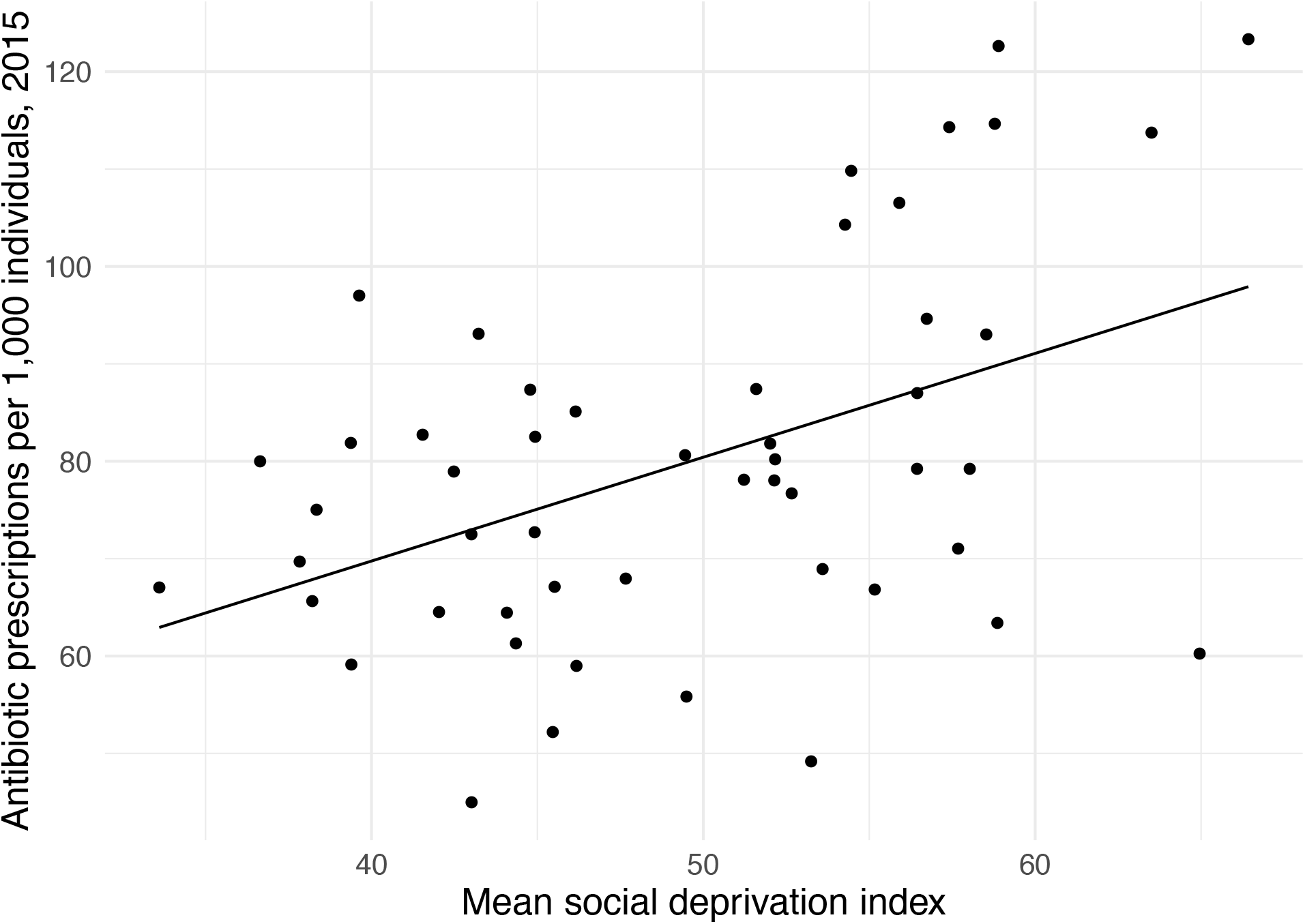
Antibiotic prescriptions per 1,000 individuals in 2016 *vs*. the mean social deprivation index (SDI) by US state. The mean SDI was calculated as the average SDI across all census tracts within a state. At the state level, there is a positive relationship (slope *p*-value < 0.001).

**Table S1.** Sex- and age-adjusted Poisson regression coefficients and 95% confidence intervals for three predictors of outpatient visit rates per 1,000 individuals at the census tract level and three medical conditions within Boston (see Table 3 in the main text for the regressions including all of Massachusetts). The vaccination rate of kindergarteners is not included here because that value was reported at the county level and therefore was constant across all Boston census tracts.

|  | Sinusitis | Pharyngitis | Suppurative otitis media |
| --- | --- | --- | --- |
| **Fraction of population under 10** | -2.20<br>(-2.37, -2.04) | -0.439<br>(-0.558, -0.320) | -1.40<br>(-1.57, -1.24) |
| **Average PM2.5** | -1.24<br>(-1.34, -1.14) | -1.41<br>(-1.48, -1.34) | -1.50<br>(-1.60, -1.41) |
| **Social deprivation index** | -1.26<br>(-1.29, -1.23) | -1.03<br>(-1.05, -1.00) | -1.20<br>(-1.23, -1.18) |

## Notes

### Competing Interest Statement

The authors have declared no competing interest.

### Funding Statement

The authors received no funding for this work.

